# Do slowly expanding lesions correspond to chronic active multiple sclerosis lesions? An integrated imaging analysis study

**DOI:** 10.64898/2025.12.03.25341192

**Authors:** Colin Vanden Bulcke, Anna Stölting, Serena Borrelli, Benoît Macq, Meritxell Bach Cuadra, Martina Absinta, Pietro Maggi

## Abstract

Chronic active lesions (CALs) are a hallmark of multiple sclerosis (MS) pathology, associated with extensive tissue damage, disability progression, and overall disease burden. Histopathologically, they consist of a hypocellular core surrounded by a rim of iron-laden, chronically activated microglia/macrophages. Proposed magnetic resonance imaging (MRI) biomarkers of CALs are paramagnetic rim lesions (PRLs) or slowly expanding lesions (SELs), detected respectively on susceptibility-based images or on longitudinal conventional MRI. While PRLs are histopathologically validated *in vivo* correlates of CALs, SELs lack pathological validation. In this study, we examine the relationship between SELs and PRLs and evaluate the robustness of the SEL detection algorithm in 56 MS participants, all imaged using a strictly homogeneous protocol on the same 3T scanner for three consecutive timepoints. PRLs included distinct subgroups of both shrinking and expanding lesions (p < 0.001), challenging the assumption that CALs necessarily expand over time. SEL detection demonstrated instability across different input resolution and segmentation methods, yielding a mean dice similarity score of 0.375. In random forest analysis, SEL volume showed inconsistent and weaker predictive value for MS disability and severity (EDSS, MSSS) compared to PRL volume. Critically, lesion-level overlap between SELs and PRLs was negligible (Cohen’s κ = –0.022) once corrected for overlap by chance. Taken together, our findings indicate that SELs and PRLs are essentially independent, underscoring the need for a refined longitudinal volumetric definition to establish a reliable CALs’ biomarker.

## 1. Introduction

Multiple sclerosis (MS) is the most prevalent chronic inflammatory disease of the central nervous system (CNS). It is characterized by recurrent neurological symptoms and the formation of focal demyelinating lesions in the brain and spinal cord (Reich et al., 2018). In their early stage, acute inflammatory MS lesions are marked by blood–brain barrier disruption (Katz et al., 1993), a key feature associated with MS relapses and predictive of short-term disease outcomes (Smith et al., 1993). After this acute phase resolves, newly formed MS lesions may evolve into either chronic active lesions (CALs) (Prineas et al., 2001) or chronic inactive lesions, the latter showing variable degrees of remyelination (Prineas & Connell, 1979).

CALs represent focal areas of compartmentalized inflammation, confined within the CNS behind a relatively intact blood–brain barrier (Lee et al., 2018). Histopathologically, CALs are typically defined by a hypocellular core containing few macrophages or microglia, surrounded by a peripheral rim of iron-laden, chronically activated microglia/macrophages (Absinta et al., 2016). Both *ex vivo* and *in vivo* studies have demonstrated that CALs are associated with extensive tissue damage (Maggi et al., 2021; Vanden Bulcke et al., 2024) and are considered strong predictors of disease severity (Absinta et al., 2019; Hemond et al., 2022; Maggi et al., 2020, 2023).

Two principal approaches are currently accepted for identifying CALs *in vivo* using magnetic resonance imaging (MRI): (i) paramagnetic rim lesions (PRLs) (Absinta et al., 2016), detected on optimized susceptibility-sensitive MRI sequences (Sati et al., 2014), and (ii) slowly expanding lesions (SELs), defined on longitudinal clinical T1-weighted (T1w) and T2-weighted (T2w) MRI (Elliott et al., 2019). Existing MRI-histopathological evidence supports the identification of PRLs as the *in vivo* correlate of histopathologically defined CALs (Bagnato et al., 2024). However, the sensitivity and specificity of PRL detection can vary across studies, and are affected by multiple factors, including the density of iron+ cells, the specific type of MRI sequence used, the MRI field strength, as well as the rater’s expertise. To mitigate this variability, rigorous and conservative PRL detection guidelines have been recently established (Bagnato et al., 2024), and this biomarker has now been proposed for the routine clinical diagnostic work-up of MS patients (Montalban et al., 2025).

In contrast, SELs are characterized by concentric, gradual expansion of chronic MS lesions (Elliott et al., 2019). Their association with disability progression (Calvi, Carrasco, et al., 2022; Preziosa et al., 2022; Wood, 2022) and ongoing tissue damage (Calvi, Carrasco, et al., 2022; Elliott et al., 2019) has led to the hypothesis that SELs may also serve as an *in vivo* surrogate of CALs (Calvi, Carrasco, et al., 2022; Elliott et al., 2019). Unfortunately, direct MRI-histopathological validation of SELs is not feasible due to their inherently longitudinal imaging definition. Moreover, only a limited number of studies have investigated the relationship between SELs and PRLs, reporting at best a moderate overlap between these two biomarkers (Calvi, Clarke, et al., 2022; Elliott et al., 2023). This highlights a persistent knowledge gap regarding the pathological validation of SELs as markers of CALs, as recently highlighted in the North American Imaging in Multiple Sclerosis (NAIMS) cooperative consensus statement (Bagnato et al., 2024).

The objectives of this study are to: (i) evaluate the longitudinal volumetric and microstructural dynamics in PRLs, (ii) assess the robustness of SEL identification against input data heterogeneity, and (iii) investigate the relationship between SELs and PRLs.

## 2. Materials and methods

### 2.1. Participants

Imaging and clinical data were consecutively collected under an institutional review board-approved protocol in adults with a diagnosis of MS followed at Cliniques Universitaires Saint-Luc (CUSL). We performed (i) a longitudinal study to investigate SEL in 56 MS patients (46 relapsing remitting (RRMS) and 10 primary or secondary progressive (PMS) with available 3 longitudinal/yearly MRI timepoints (see below); and, (ii) a cross-sectional study performed in 73 MS patients (54 RRMS and 19 PMS) in order to compare the performance of the different segmentation tools tested as variable inputs for the SEL detection algorithm. All patients were (1) ≥18 years old, had (2) an MS diagnosis according to the 2024 McDonald criteria (Montalban et al., 2025), (3) and a clinical expanded disability status state (EDSS) and multiple sclerosis severity score (MSSS) evaluation at baseline. The longitudinally homogeneous 3-Tesla (3T) MRI protocol for the longitudinal study included a 3-dimensional (3D) T2*-weighted echo-planar imaging (EPI) (Sati et al., 2014) (for PRL assessment), T2-weighted fluid-attenuated inversion recovery (FLAIR) and T1-weighted magnetization prepared rapid gradient echo image (MPRAGE) (for lesion segmentation and SEL assessment), and a quantitative T1-map derived from a sagittal 3D magnetization-prepared 2 rapid gradient echo (MP2RAGE) (for microstructural investigations). Patients included in the cross-sectional study had available FLAIR and MPRAGE images, and a whole-brain manual lesion segmentation by an experienced rater (AS). This study was approved by the local ethical committee of CUSL, and all participants provided written informed consent according to the declaration of Helsinki prior to the study.

### 2.2. MRI imaging protocol

MRI acquisition was performed on a 3T whole-body MR scanner (GE SIGNA^TM^ Premier research scanner, General Electric, Milwaukee, WI) equipped with a 48-channel head coil. The MRI protocol included: (i) sagittal 3D MPRAGE sequence (repetition time (TR)=2186 ms, echo-time (TE)=3 ms, inversion time (TI)=900 ms, field of view (FOV)=256 mm, number of slices (#slices)=156, voxel size=1.0×1.0×1.0 mm), (ii) sagittal 3D FLAIR sequence (TR=5000 ms, TE=105 ms, TI=1532 ms, FOV=256 mm, #slices=170, voxel size=1.0×1.0×1.0 mm), (iii) a sagittal high-resolution 3D EPI (Sati et al., 2014) sequence (TR=80.2 ms, TE=35 ms, flip angle=18°, FOV=256 mm, #slices=355, voxel size=0.67×0.67×0.67 mm), (iv) quantitative T1-map derived from a sagittal 3D MP2RAGE sequence (TR=1925 ms, TE=3 ms, TI=700 and 2500 ms, FOV=256 mm, #slices=156, voxel size=1.0×1.0×1.0 mm), and (v) a post-gadolinium T1-weighted spoiled gradient recalled echo (CE-SPGR) sequence (TR*=*6.9 ms, TE*=*2.1 ms, flip angle=12°, FOV=240, #slices=150, voxel size=0.69×0.69×1.0 mm).

### 2.3. Slowly expanding lesions algorithm

This section describes the implementation of the SEL detection algorithm and outlines the experiments conducted to test its underlying assumptions, evaluate the robustness of its implementation, and evaluate PRL-SEL co-localization.

#### 2.3.1. Longitudinal volume assessment of PRLs

The detection of SELs relies on the assumption that CALs expand concentrically-outward over time due to the presence of persistent smoldering inflammation at their edges. This lesional expansion can be identified *in vivo* using clinically available MRI sequences. To test this assumption, we systematically screened our cohort to identify all PRLs at baseline, in order to investigate their longitudinal volumetric and microstructural behavior.

For this purpose, 3D FLAIR and CE-SPGR images were rigidly coregistered using mutual information to the high-resolution 3D EPI scan using ANTs (Avants et al., 2008). Chronic non-enhancing MS lesions were classified as PRL if they showed no enhancement on post-gadolinium CE-SPGR images and demonstrated a paramagnetic rim on unwrapped filtered phase images (Absinta et al., 2013), in accordance with the latest consensus criteria (Bagnato et al., 2024) (see Figure 1). PRL identification required the following features, as previously described (Maggi et al., 2023): (i) colocalization with the edge of an MS lesion on FLAIR, (ii) visibility in at least two planes, and (iii) a paramagnetic rim covering at least two-thirds of the lesion’s white matter border on the slice of maximum visibility. PRL assessment was performed by consensus of two trained raters (AS and PM).

**Figure 1.**
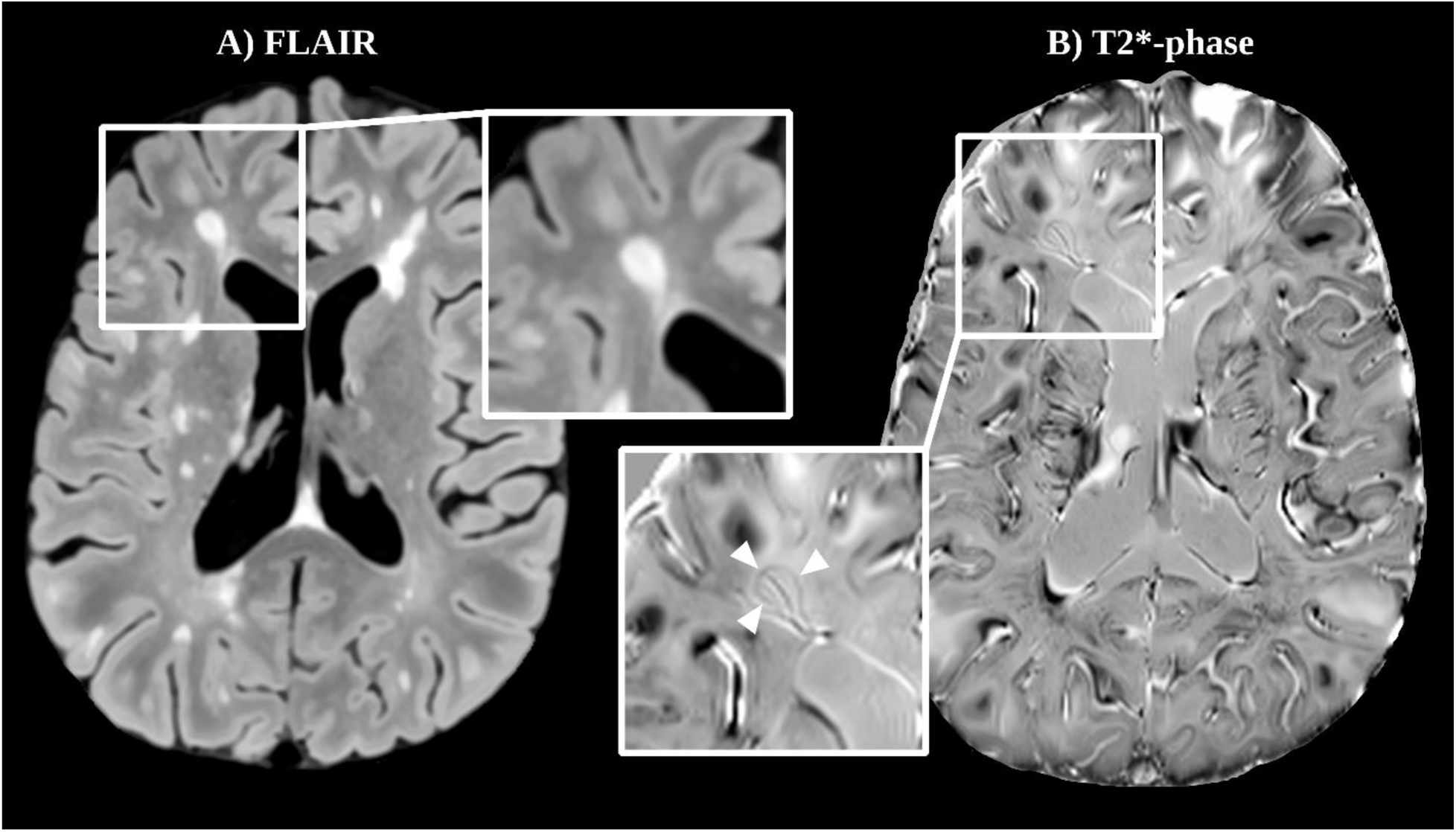
Axial fluid-attenuated inversion recovery (FLAIR — A) and T2*-weighted unwrapped-filtered phase (B) images of an individual in their 20’s with relapsing-remitting multiple sclerosis (RRMS) showing a typical example of a paramagnetic rim lesion (PRL — white arrowheads).

Whole-brain lesion segmentation was performed using FLAMeS (Dereskewicz et al., 2025; Isensee et al., 2021) applied to the FLAIR image for semantic binary segmentation, combined with an in-house algorithm to track longitudinal lesion volume changes (detailed in supplementary materials). Lesion volume trajectories were quantified by fitting a Huber (Huber & Ronchetti, 1980; Owen, 2007) linear regression model to normalized annual lesion volume changes. Lesions were classified as expanding if the slope exceeded +9%, shrinking if below – 9%, or stable if between –4% and +4%, based on the distribution of our data. Lesions not meeting the ± 9% criteria were defined as having a tendency to expand (or shrink). Finally, longitudinal PRL and non-PRL volume dynamics were assessed using a repeated-measures linear mixed-effects model of log-transformed lesion volume over time, including a random subject and lesion effect. In addition, the microstructural evolution of mean lesion T1 values was investigated using repeated-measures linear mixed-effects models (with random intercepts for subjects and lesions), to test the effects of time and its interactions with PRL status (PRL, non-PRL), lesion volume evolution (shrinking, expanding, stable), and their combination. Pairwise comparisons were performed using Tukey-adjusted post-hoc tests (emmeans), and the corresponding contrast estimates (β) and confidence intervals are reported.

#### 2.3.2. Implementation of SEL detection

The SEL detection algorithm (Figure 2) implemented in this work was designed as a faithful re-implementation of the original method described by Elliott et al. (Elliott et al., 2019). Since the original algorithm is not open-source, certain methodological aspects required interpretative implementation choices based on the descriptions provided in the publication. The final implementation has been released as an open-source BMAT-App within the BMAT software (Vanden Bulcke et al., 2022).

**Figure 2.**
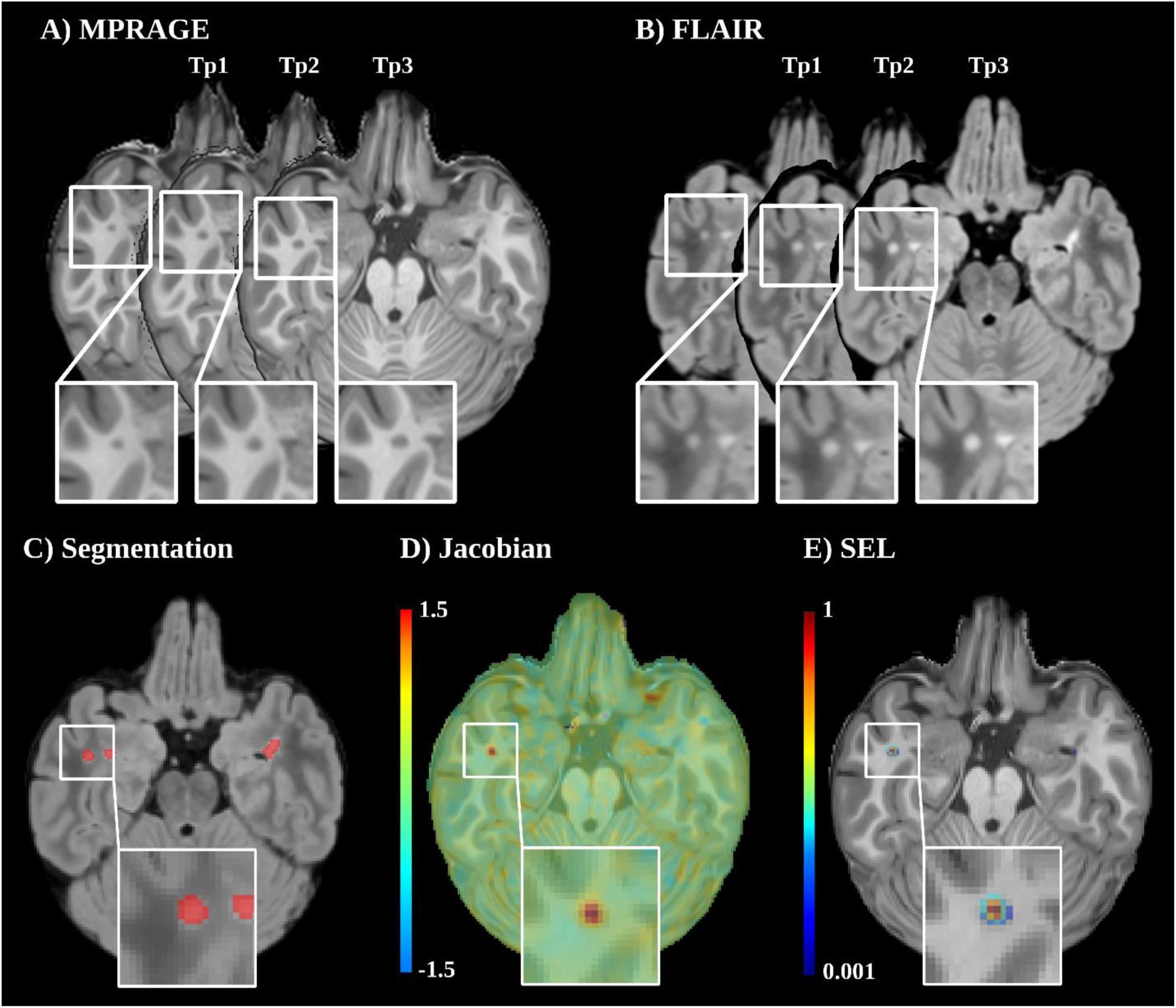
Representation of a slowly expanding lesion (SEL) found in an individual in their 20’s with relapsing-remitting multiple sclerosis (RRMS). The SEL is detected using a Jacobian-based analysis of the deformation field computed via a non-rigid registration between (A) magnetization prepared rapid gradient echo (MPRAGE) and (B) fluid-attenuated inversion recovery (FLAIR) images of three different timepoints (Tp1, Tp2, Tp3). (C) represents an automatic binary lesion segmentation, which is combined with the Jacobian determinant of the deformation field between Tp1 and Tp3 (D), to identify SEL (E; here displayed as an overlay between the MPRAGE image and the Jacobian map).

The algorithm requires FLAIR and MPRAGE images as inputs acquired at three timepoints per subject (Figure 2.A and B). Both modalities are first denoised using a non-local means adaptive filter (Manjón et al., 2010), and skull-stripped with SynthStrip (Hoopes et al., 2022). Finally, the FLAIR image is rigidly registered using mutual information to the MPRAGE space using ANTs (Avants et al., 2008). Following preprocessing, deformation fields are computed between the baseline (first timepoint) and the subsequent second and third timepoints. To compute this deformation field, the MPRAGE images from each pair of timepoints are registered in a halfway space using *mri_robust_register* from FreeSurfer (Reuter et al., 2010) with the dedicated halfway option. The corresponding FLAIR images and brain masks are then transformed into the same halfway space using the resulting transformation matrix. A non-rigid diffeomorphic registration algorithm from ANTs (Avants et al., 2008) (step size = 0.7; Gaussian sigma = 2) is subsequently applied to the MPRAGE and FLAIR image pairs in the halfway space to compute the deformation field between two timepoints. Finally, the Jacobian determinant of the deformation field is calculated with ANTs (Avants et al., 2008), resulting in a scalar map that quantifies local expansion and shrinkage (Figure 2.D).

SEL candidates are identified using a binary lesion segmentation (Figure 2.C) in combination with the Jacobian determinant of the deformation field between the first and last timepoints. The Jacobian map is first normalized to a one-year interval. A SEL candidate (Figure 2.E) is then defined as a connected region within the lesion mask exhibiting a peak local expansion of at least 12.5%, surrounded by adjacent voxels showing a minimum of 4% expansion (Elliott et al., 2019), as determined using a hysteresis thresholding approach (Canny, 1986). If multiple clusters of local expansion fulfilling these criteria occur within the same connected lesion area, each cluster is treated as an independent SEL candidate, consistent with previous methodology (Preziosa et al., 2022).

As originally described by Elliott et al. (Elliott et al., 2019), SEL candidates are then defined as definitive SELs based on two supplementary criteria: *concentricity* and *constancy* criteria. Concentricity was defined as an inside-out expansion pattern (Figure 2.E), where the Jacobian determinant is greatest at the center of a SEL candidate and decreases towards the edge. Quantitatively, the SEL candidate mask was subdivided into concentric voxel-wise bands, and concentricity was measured as the slope of the best Huber (Huber & Ronchetti, 1980; Owen, 2007) linear regression of mean Jacobian values in each concentric band as a function of radial distance from the mask edge. Constancy was defined as the longitudinal stability of lesion expansion across different timepoints. Specifically, for each SEL candidate, the mean Jacobian-derived expansion was modeled as a Huber (Huber & Ronchetti, 1980; Owen, 2007) linear regression over time, and constancy was quantified as the mean normalized squared residual error between the observed expansion values and the fitted linear trajectory, with lower residuals indicating more constant expansion over time. The concentricity and constancy metrics were then standardized into z-scores compared to the SEL candidates cohort and combined into a single scalar score S, defined as the difference between the z-score normalized concentricity and the z-score normalized constancy. Finally, a SEL candidate is defined as definite SEL (Figure 2.E) if its scalar score S ≥ 0. SEL candidates with a score S < 0 are considered as possible SEL (SELpos).

#### 2.3.3. Evaluation of the SEL detection algorithm

In this study, in order to assess the reproducibility and stability of the SEL detection algorithm relative to its inputs, we systematically evaluated its performance under varying imaging and segmentation conditions. First, to simulate a routine clinical imaging protocol, the isotropic 1 mm^3^ FLAIR image was degraded to a 1×1×3 mm axial FLAIR and subsequently resampled back to 1 mm^3^ isotropic resolution, as originally described by Elliott et al. (Elliott et al., 2019). Hereafter, this condition is referred to as *2D* throughout the paper, while the original isotropic image is referred to as *3D*. For both *2D* and *3D* images, binary lesion segmentations were generated using either FLAMeS (Dereskewicz et al., 2025) or SAMSEG (Cerri et al., 2021) (referred to as _flames or *_samseg*), in order to evaluate the robustness of the SEL detection algorithm across lesion segmentation methods of varying performance.

The segmentation performance of FLAMeS- and SAMSEG-derived segmentations was evaluated against an expert manual segmentation (AS) using the Dice Similarity Coefficient (DSC) (Dereskewicz et al., 2025), and group-level differences were assessed using a Mann–Whitney U test. To compare the effect of segmentation and image resolution on SEL detection, we performed an ANOVA on the number and volume of SELs at the subject level. The spatial overlap of SEL masks across conditions (2D vs. 3D; FLAMeS vs. SAMSEG) was further quantified using the DSC.

#### 2.3.4. Association with clinical disability

The association between clinical disability scores and SEL burden was investigated by fitting linear regression models with EDSS as the outcome and log-transformed SEL volume (log_vSEL) as the predictor, adjusting for baseline age, sex assigned at birth, and brain volume at baseline (computed using FreeSurfer) (Fischl, 2012). To investigate lesion-specific tissue characteristics, we compared T1 values between SEL and non-SEL lesions at baseline, as well as longitudinal T1 changes between baseline and the third MRI timepoint, using a linear mixed-effects model with a random subject and lesion intercept. Finally, to evaluate the predictive value of SEL-derived volume for disability measures, we performed a random forest analysis (R, *randomForest* package) (Liaw & Wiener, 2007) with 10-fold cross-validation, using EDSS and MSSS as outcome variables. Candidate predictors included normalized values (scaled 0–1) of log_vSEL_3D_flames, log_vSEL_2D_flames, log_vSEL_3D_samseg, log_vSEL_2D_samseg, brain volume, log_vLesions, log_vPRL, age (only for EDSS prediction), and sex.

#### 2.3.5. Lesion-level assessment of PRL-SEL overlap

Lastly, spatial lesion-level overlap between PRLs and SELs was assessed. PRLs were manually and systematically assessed by two expert raters (AS and PM), as detailed in the *Longitudinal volume assessment of PRLs* section. For each tested input condition (described above), we recorded at the subject level: (i) the total number of SELs, (ii) the number of PRLs located within the binary lesion segmentation mask of the condition (to ensure methodological consistency), and (iii) the number of overlapping SEL – PRL pairs. From these counts, we computed lesion-level prevalence values for SELs (*P(SEL)*) and PRLs (*P(PRL)*), as well as the conditional probability (*P(SEL|PRL)* & *P(PRL|SEL)*) across the cohort. To more rigorously characterize the relationship between SELs and PRLs, we further derived four complementary overlap metrics, designed to account for the overlap expected by chance under the assumption of two independent distributions drawn from the same lesion population:

- **Joint probability:** P(*SEL* ∩ *PRL*) = P(*PRL*) ⋅ *P*(*SEL* |*PRL*) = P(*SEL*) ⋅ *P*(*PRL*|*SEL*), representing the probability that a lesion is classified as both a SEL and a PRL.
- **Cohen’s Kappa (κ)**: 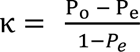, where P is the observed agreement and P_e_ is the agreement expected by chance; 0 indicates no agreement beyond chance, and 1 indicates perfect agreement between the two biomarkers.
- **Jaccard Index (JI)**: 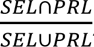, quantifying the proportion of shared lesions between SELs and PRLs relative to the total number of unique lesions identified by either biomarker.
- **Mutual Information (MI)**: 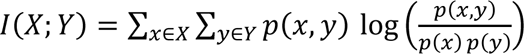, quantifying the amount of shared information between SEL and PRL biomarkers, with higher values indicating greater correspondence.

As a supplementary analysis, we conducted a re-analysis of the two previous studies that have explored the overlap between SELs and PRLs (Calvi, Clarke, et al., 2022; Elliott et al., 2023). While both studies have reported the raw overlap between these two MRI biomarkers, they did not compute any further agreement metrics. Therefore, we propose to re-analyze the results of these studies with the same framework described above. Since the second study from Elliott et al. (Elliott et al., 2023) did not report the total number of lesions in their study, thus preventing us from computing the lesion-level biomarker prevalence, we used values computed in external studies for the prevalence of PRL (Ng Kee Kwong et al., 2021) and prevalence of SEL (Calvi, Carrasco, et al., 2022), introducing a certain bias to take into consideration.

## 3. Results

### 3.1. Demographics

Fifty-six participants were included in the SEL detection experiment. The baseline demographics, clinical and MRI characteristics are reported in Table 1. The median EDSS and MSSS at baseline were 2 (0 – 7) and 5.26 (0.62 – 9.46) respectively. Thirty-seven out of fifty-six (66.1%) of participants had at least one PRL and the mean total PRL volume was 1840.23 mm^3^.

**Table 1.** Demographics, clinical and MRI characteristics of the participants included in the SEL detection experiment.

| Demographics and clinical metrics | Values |
| --- | --- |
| Number of patients (RMS, PMS) | 56 (46, 10) |
| Age, mean (range) [years] | 42.36 (19 – 72) |
| Female, N (%) | 36 (64.3%) |
| Baseline disease duration, mean (range) [years] | 7.98 (0 – 37.24) |
| EDSS at baseline, median (range) | 2 (0 – 7) |
| MSSS at baseline, median (range) | 5.26 (0.62 – 9.46) |
| Normalized brain volume, mean ± std | 0.745 ± 0.029 |
| Normalized gray matter volume, mean ± std | 0.413 ± 0.026 |
| Normalized white matter volume, mean ± std | 0.243 ± 0.021 |
| Number of PRL, mean (range) | 5.84 (0 – 91) |
| PRL volume, mean ± std [mm<sup>3</sup>] | 1840.23 ± 4521.49 |
| $\geq 1$ PRL, N (%) | 37 (66.1%) |

### 3.2. Longitudinal volume assessment of PRLs

Out of the 498 PRLs identified at baseline, 361 (72.5%) were classified as shrinking, 51 (10.3%) as expanding, and 46 (9.2%) as stable. Among the remaining lesions, 24 (4.8%) demonstrated a trend toward shrinkage and 16 (3.2%) a trend toward expansion, though neither group met the predefined 9% classification threshold. Group analyses confirmed that shrinking PRLs continued to significantly decrease in volume over time (β=-1.37e-04 [95% CI: -1.70e-04; -1.04e-04], p < 0.001), whereas expanding PRLs continued to increase in volume (β=2.48e-04 [95% CI: 1.19e-04; 3.77e-04], p < 0.001). Moreover, shrinking PRLs displayed a significantly different volumetric trajectory compared to expanding PRLs (p < 0.001). Taken all together, PRLs exhibited a significant longitudinal decrease in log-transformed volume (β=-7.88e-05 [95% CI: -1.10e-04; - 4.78e-05], p < 0.001).

Similarly, non-PRL lesions also demonstrated a significant overall tendency to shrink in log-transformed volume over time (β=-4.12e-05 [95% CI: -5.56e-05; -2.69e-05], p < 0.001). Within this group, subsets of lesions were identified as shrinking, expanding, or stable, each showing significant longitudinal trends consistent with their classification (p < 0.001) and different from the other sub-groups (p < 0.001).

Regarding the microstructural evolution of T1, PRLs exhibited higher baseline T1 values than non-PRLs (β=+165 ms [95% CI: +143.64; +186.36], p < 0.001), as well as a stable T1 evolution over time (β=-0.0152 [95% CI: -0.0482; 0.0177], p=0.36) compared to a significant decrease in T1 for non-PRL (β=-0.1497 [95% CI: -0.1643; -0.1352], p < 0.001). No significant T1 differences were observed at baseline between shrinking, stable, and expanding lesions; however, shrinking lesions exhibited a significantly (p < 0.05) steeper T1 decline (β=-0.160 [95% CI: - 0.177; -0.1435], p < 0.001) compared to expanding lesions (β=-0.1 [95% CI: -0.139; -0.0606], p = 0.203). When considering the interaction between PRL status and lesion volume trajectory, only shrinking lesions showed a differential T1 evolution with regards to PRL status, with shrinking PRLs demonstrating a T1 increase over time (β=0.017 [95% CI: -0.018; 0.053]), significantly different (p < 0.001) from shrinking non-PRLs exhibiting a T1 decrease (β=-0.211 [95% CI: -0.229; -0.192], p < 0.001). No significant T1 changes were observed across PRL subgroups (shrinking, stable, or expanding; all p > 0.05). In contrast, shrinking non-PRLs showed a significantly steeper decrease in T1 compared to expanding non-PRLs (p < 0.001).

### 3.3. Evaluation of SEL detection

At baseline, FLAMeS segmentation outperformed SAMSEG, with higher overlap against the manual reference standard (DSC = 0.54 ± 0.16 vs. 0.45 ± 0.15, p < 0.01).

When comparing SEL detection across different input segmentations and image dimensions at a lesion-level, DSC between SEL masks was weak to moderate but comparable across segmentation methods and input resolution. More specifically, 3D_flames versus 2D_flames achieved a DSC of 0.37 ± 0.28, while 3D_samseg versus 2D_samseg yielded a DSC of 0.36 ± 0.30. When comparing FLAMeS to SAMSEG, the DSC was 0.41 ± 0.32 for 3D and 0.36 ± 0.32 for 2D. At a subject-level in contrast, minor differences were observed with the ANOVA analysis, as presented in Table 2. The number of SELs per subject ranged from 2.57 (2D_samseg) to 3.50 (2D_flames), with significantly more SELs detected using 2D_flames than 2D_samseg (p < 0.001). Total SEL volume per subject ranged from 69.48 mm³ (3D_flames) to 102.48 mm³ (2D_flames), with significantly greater volumes detected in 2D_flames compared to 3D_flames (p < 0.05). The rest of the pairwise comparisons were non-significant.

**Table 2.** SEL-derived metrics at a subject level computed for the different inputs images 3D vs 2D FLAIR) and segmentation (FLAMeS vs SAMSEG).

|  | 3D_flames | 3D_samseg | 2D_flames | 2D_samseg |
| --- | --- | --- | --- | --- |
| # T2 lesions, N (range) | 43.36 (3 – 186) | 28.18 (3 – 97) | 28.7 (3 – 116) | 20.29 (3 – 69) |
| # nonSEL, N (range) | 37.20 (2 – 185) | 24.43 (2 – 97) | 23.70 (1 – 115) | 17.55 (2 – 68) |
| # SELpos, N (range) | 11.32 (1 – 69) | 7.68 (0 – 51) | 10.09 (0 – 47) | 8.29 (0 – 37) |
| # SEL, N (ranges) | 3.16 (0 – 16) | 2.77 (0 – 10) | 3.5 (0 – 16) | 2.57 (0 – 13) |
| PSEL, % | 7.29% | 10.19% | 12.20% | 12.67% |
| vSEL, mean ± std (mm<sup>3</sup>) | 69.48 ± 113.87 | 82.36 ± 106.48 | 102.48 ± 145.99 | 83.61 ± 120 |
| $\geq 1$ SEL, N (%) | 41/56 (73.21%) | 43/56 (76.79%) | 50/56 (89.29%) | 44/56 (78.57%) |

At the lesion level, SELs showed a non-significant trend toward higher T1 values compared to non-SELs. However, SELs exhibited a greater increase in T1 between baseline and last follow-up, significant for 3D_flames (p < 0.001), 3D_samseg (p < 0.05), and 2D_samseg (p < 0.05), but not for 2D_flames (p = 0.106).

### 3.4. Association with clinical disability

Overall, in our cohort, no significant associations were observed between log-transformed SEL volume (log_vSEL) and EDSS. Random forest modeling identified brain volume, age, log-transformed lesion volume (log_vLesions), and log_vPRL as the variables with the strongest predictive value in explaining EDSS (see Figure 3.A). More specifically, log_vSEL_3D_flames and log_vSEL_2D_flames showed moderate to weak importance, about 40% and 74% less important than log_vPRL respectively. Specifically, predictive performance of log_vSEL varied substantially depending on input type, ranging from 3.499 in %IncMSE (log_vSEL_3D_flames) to - 2.84 %IncMSE (log_vSEL_3D_samseg), the latter even showing no predictive value/importance. Regarding MSSS, log_vLesions, log_vPRL, and brain_volume were the strongest predictors (see Figure 3.B). Once more, log_vSEL predictive values varied substantially depending on inputs, with a reduction of about 30% and 59% of importance for log_vSEL_2D_flames and log_vSEL_3D_flames when compared to log_vPRL, and with log_vSEL_3D_samseg showing again no predictive value of disease severity.

**Figure 3.**
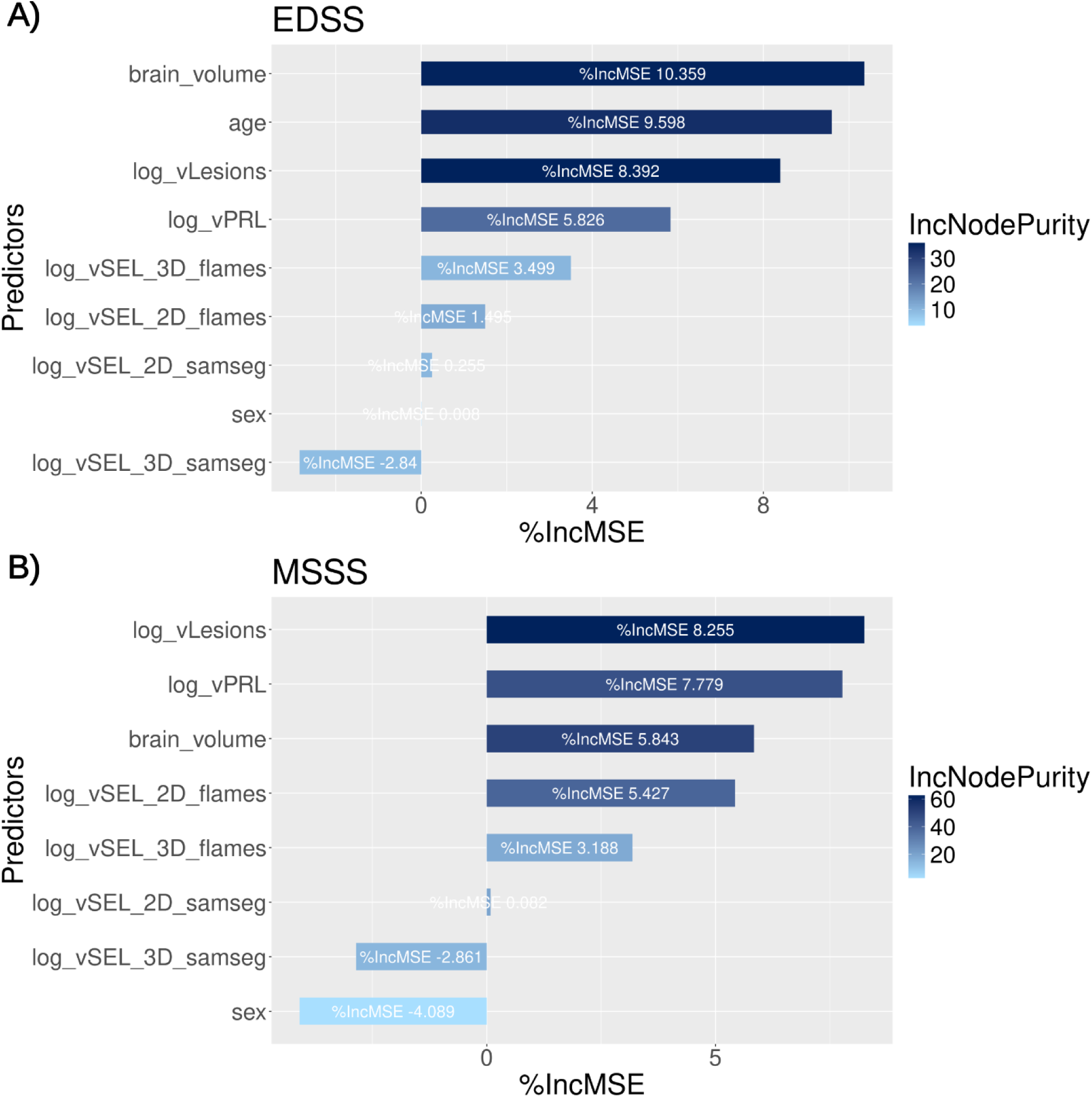
Random Forest analysis of the predictive value of different imaging and clinical values on expanded disability status scale (EDSS, A) and on multiple sclerosis severity score (MSSS). The predictors are normalized brain volume, age, sex, log-transformed total T2 lesion volume (log_vLesions), PRL volume (log_vPRL), SEL_3D_flames volume (log_vSEL_3D_flames), SEL_3D_samseg volume (log_vSEL_3D_samseg), SEL_2D_flames volume (log_vSEL_2D_flames), SEL_2D_samseg volume (log_vSEL_2D_samseg).

### 3.5. Lesion-level assessment of PRL-SEL overlap

Finally, table 3 shows the lesion-level overlap of SELs against PRLs under the different input conditions. The mean prevalence of SELs across inputs was 9.11%, compared to 19.65% for PRLs. The conditional probability of a lesion being a SEL given it was a PRL was 7.73%, whereas the probability of a lesion being a PRL given it was a SEL was 16.85%. Overall lesion-level agreement between SELs and PRLs was essentially nonexistent (P(*SEL* ∩ *PRL*) = 1.57%, Cohen’s κ = –0.022; JI = 0.056; MI = 0.00079).

**Table 3.**
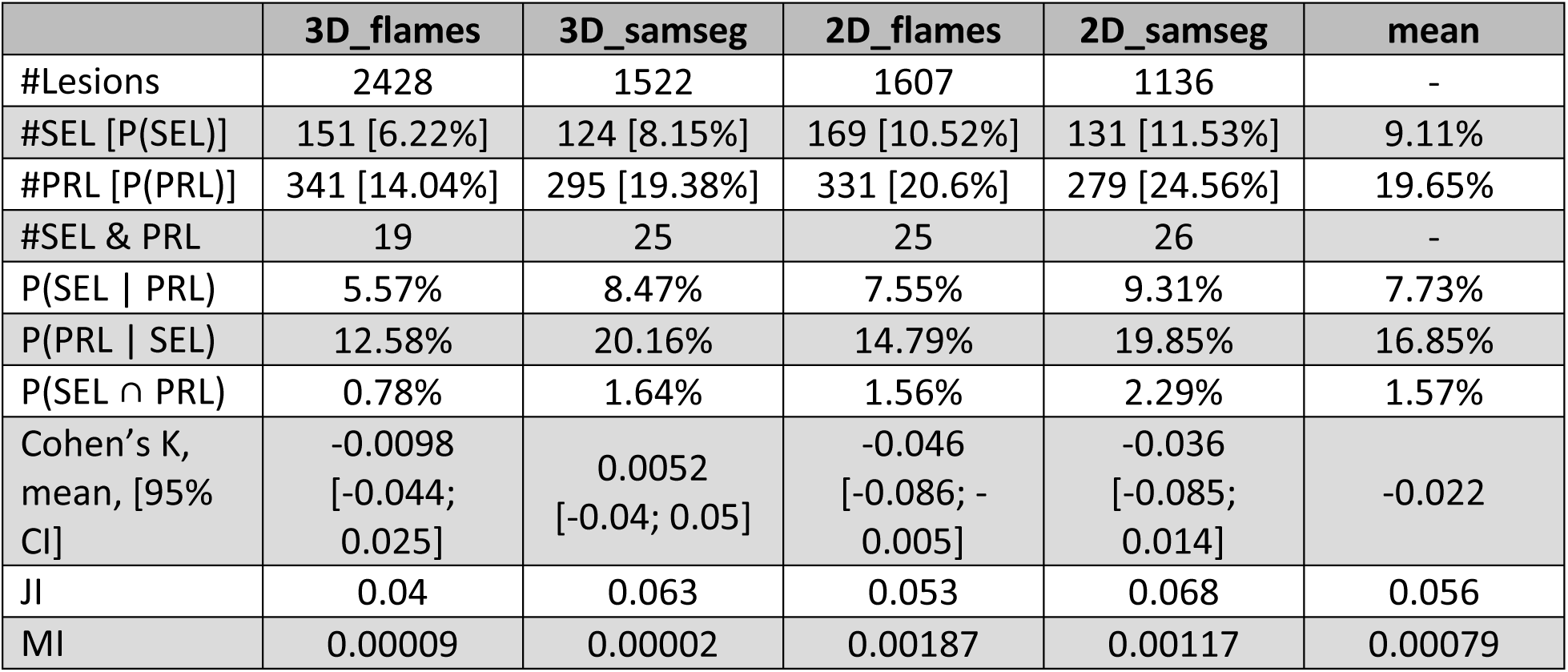
Lesion-level correspondence of SELs, computed with different input images (3D vs 2D FLAIR) and segmentation (FLAMeS vs SAMSEG), and PRLs. Abbreviations: prevalence of SEL [P(SEL)], prevalence of PRL [P(PRL)], conditional probability of being a SEL if a lesion is a PRL [P(SEL | PRL)], conditional probability of being a SEL if a lesion is a PRL [P(PRL | SEL)], joint probability of SEL and PRL [P(SEL ∩ PRL)], Cohen’s agreement Kappa (Cohen’s K), jaccard index (JI), and mutual information (MI), 95% confidence interval [95% CI].

The first study from Calvi et al. (Calvi, Clarke, et al., 2022) reported these results: out of 1492 lesions, 616 were SELs, 80 were PRLs, and 43 were both PRLs and SELs. Re-analyzing these data using the same framework as our study yields a joint probability of 2.9%, a Cohen’s kappa of – 0.04, a Jaccard index of 0.066, and a mutual information of 0.004 — all results consistent with our study. The second study, from Elliott et al. (Elliott et al., 2023), reported that 47/119 PRLs (39.5%) corresponded to SELs while 46/267 (17.23%) SELs corresponded to PRLs, but they did not report the total number of lesions in their cohort, preventing a direct prevalence-based calculation. Using external prevalence estimates (29% for SELs (Calvi, Carrasco, et al., 2022) and 9.8% for PRLs (Ng Kee Kwong et al., 2021)), we obtain a joint probability of 11.5%, a Cohen’s kappa of 0.52, a Jaccard index of 0.421, and mutual information of 0.21 — suggesting a moderate overlap. However, using prevalences from different studies undermines the validity of this estimate as the prevalence of these biomarkers varies substantially between studies. More specifically, the computed conditional probability P(SEL | PRL) based on the computed joint probability of 11.5% and P(PRL) of 9.8% exceeded 100% (∼117%), an impossibility that indicates a sizeable overestimation of these computed overlap metrics.

## 4. Discussion

The objective of this study was to evaluate the SEL detection algorithm, assessing its consistency across heterogeneous imaging inputs (including variation in image resolution and lesion segmentation) and disentangling the relationship between SELs and PRLs, the latter representing a pathologically validated biomarker of CALs.

### 4.1. Rethinking volumetric dynamics of CALs

The SEL algorithm aims to identify concentric, gradual expansion of chronic MS lesions using clinically available MRI sequences (Elliott et al., 2019). The core assumption of SEL as a biomarker of CAL is that CALs would expand concentrically over time due to persistent smoldering inflammation at their edges. While this idea is conceptually plausible, it likely oversimplifies the underlying MS lesions’ pathophysiology and is increasingly challenged by emerging *in vivo* evidence (Sethi et al., 2017). Direct histopathological validation is not feasible, since lesion expansion cannot be assessed post-mortem, leaving longitudinal MRI as the only available tool for this evaluation.

Following the resolution of gadolinium enhancement (generally lasting 1-2 months), chronic MS lesions can follow different pathological trajectories, which in turn affect their short- and medium-term volumetric changes. Both remyelination and tissue loss are likely to drive lesion shrinkage, while smoldering edge inflammation with ongoing demyelination can lead to expansion. In this study, we identified distinct subsets of expanding, shrinking, and stable PRLs, confirming the heterogeneity of CALs in terms of lesional volume trajectories, and in line with previous work showing that PRLs in MS are highly heterogeneous in terms of lesional volume and tissue damage (Stölting et al., 2024). Importantly, and in agreement with previous evidence (Sethi et al., 2017), we found that contraction is the predominant pattern of MS lesions’ volumetric evolution, with the majority of chronic lesions (both PRL and non-PRL) significantly shrinking over time. From a microstructural standpoint, both PRLs and expanding lesions exhibited a more pronounced accumulation of lesional tissue damage than their respective non-PRL and shrinking counterparts, with PRL status exerting the strongest effect. Interestingly, shrinking PRLs showed a significantly greater progression of lesional tissue damage compared to shrinking non-PRLs, likely reflecting tissue loss driven by compartmentalized-inflammation-induced cell senescence (Fagiani et al., 2025). This supports the hypothesis that lesion shrinkage in chronic active and inactive lesions may be primarily driven by distinct pathological processes — likely tissue loss and remyelination, respectively (Sethi et al., 2017). In line with these observations, recent works investigating lesion volume change and quantitative tissue damage metrics have shown that shrinking lesions may display more severe baseline tissue damage than expanding ones (Ravano et al., 2024; Vanden Bulcke, 2024). All these observations critically challenge the concept of longitudinal radial expansion as a biomarker of CALs, suggesting instead that CALs undergo multiple volumetric evolution phases throughout their lifespan.

### 4.2. Limitations of SEL as a reliable biomarker of CALs

The SEL algorithm, as introduced by Elliott et al., relies on non-rigid registration to estimate local tissue deformation, specifically expansion and contraction, using Jacobian determinant analysis. A first limitation regards reproducibility; the original implementation being not publicly available, calculating SELs necessarily requires parameter re-implementation. In this study, we evaluated the robustness of the algorithm under two main sources of input heterogeneity: (i) image quality, by comparing degraded 2D FLAIR (1×1×3 mm resampled to isotropic 1 mm) with native 3D FLAIR (clinical vs research sequence), and (ii) lesion segmentation, using FLAMeS (state-of-the-art) versus SAMSEG (less performant) (Dereskewicz et al., 2025). At the subject level, the average number and total volume of SELs remained relatively consistent across input conditions. However, voxel-wise comparison revealed poor overlap between SEL masks across varying input conditions (average DSC of ∼0.375). This discrepancy suggests that while the overall SEL burden is relatively stable, the exact spatial localization of SELs is highly input-dependent, likely a consequence of the way SEL detection criteria are defined (further discussed below). At the lesion level, SELs showed a trend to higher baseline T1 and significantly greater longitudinal T1 increase between first and last timepoints, consistent with prior reports (Calvi, Carrasco, et al., 2022; Elliott et al., 2019; Preziosa et al., 2022). This effect is plausibly explained by the Jacobian-based detection being directly influenced by intensity changes in the underlying T1-weighted images. Despite these associations, in our cohort we did not observe any significant association between log-transformed SEL volumes (log_vSEL) and EDSS. Finally, in a random forest cross-validation analysis of EDSS prediction, SEL volume demonstrated moderate predictive value and its performance in predicting disability scores (EDSS and MSSS) was consistently outperformed by total T2 lesion load, and total PRLs’ volume.

Beyond the lack of robustness demonstrated in this study, several conceptual limitations exist in the original implementation of the SEL algorithm as presented by Elliott et al. (Elliott et al., 2019). A first limitation is the use of a binary lesion mask as input, rather than an instance-level lesion mask with pre-selected individual lesions. While the binary mask approach offers computational efficiency and automation, it introduces a fundamental ambiguity: SELs may not correspond to singular MS lesions as detected by MRI and potentially validated by MRI-histopathological studies. Indeed, previous studies have noted that if the same T2 hyperintense lesion includes two or more clusters of Jacobian expansion, this may reflect the presence of different lesions that became confluent (Preziosa et al., 2022), therefore using the Jacobian analysis to separate confluent lesion. However, it was acknowledged in the original paper that T1 intensity changes exclusively localized at the edge of a lesion results in a concentric inside-out Jacobian expansion pattern (more pronounce expansion in the center of the lesion compared to the edge) (Elliott et al., 2019). This illustrates that Jacobian-based expansion is not directly linked to voxelwise image intensity changes. Therefore, the hypothesis that Jacobian expansion can reliably differentiate individual lesion instances within confluent lesions lacks support.

A second major issue lies in the implementation of the concentricity and constancy criteria. Although each criterion is conceptually reasonable, their practical use introduces important limitations. Both criteria are z-score normalized against all SEL candidates in the study population, which makes SEL classification inherently population-dependent rather than lesion-specific. As a result, the very same lesion could be labeled SEL in one population but not in another. Moreover, z-score normalization enforces a distribution where a fixed proportion of lesions will have positive values, regardless of their raw criterion scores. This means that even a lesion with a raw negative concentricity value could end up with a positive z-score normalized concentricity score if the overall cohort has an even lower average. Such population-driven scaling likely explains why the number and volume of SELs at the subject level appeared rather consistent across different inputs, while lesion-level overlap remained poor. Finally, the decision to combine the normalized values of concentricity and constancy into a single abstract metric strips these measures of any biological meaning. Taken together, these limitations challenge the pathological value and utility of the SEL detection algorithm in MS. While SELs are presented as lesions with consistent concentric expansion, the actual implementation detects regions within lesion masks (not necessarily distinct lesions) using a population-normalized heuristic with no direct histopathological relevance.

### 4.3. Divergence between SELs and PRLs

A major concern with SELs is the absence of rigorous biological validation as markers of CALs. By design, SELs are purely longitudinal imaging constructs and therefore cannot be directly validated through histopathology. This makes comparisons with established *in vivo* biomarkers of CALs essential. Currently, two such biomarkers exist: PRLs, detectable with susceptibility-based MRI, and CALs identified through positron emission tomography (PET) imaging using 18-kDa translocator protein (TSPO) binding radioligands (TSPO-PET) (Bagnato et al., 2024). To our knowledge, no study has investigated lesion-level overlap between SELs and TSPO-PET-defined CALs, and only limited work has explored the correspondence between SELs and PRLs (Calvi, Clarke, et al., 2022; Elliott et al., 2023).

In our study, lesion-level correspondence between SELs and PRLs was essentially absent, with a mean joint probability of 1.57%, a mean Cohen’s kappa of –0.022, a mean Jaccard index of 0.056, and a mean mutual information of 0.00079, suggesting no meaningful relationship between PRLs and SELs. Although two other studies reported raw overlap between SELs and PRLs, neither accounted for chance-level overlap expected when comparing two biomarkers drawn from the same lesion population. Re-analyzing the available data from these studies showed similar results to our study, suggesting that SELs and PRLs are essentially independent. Given that PRLs, but not SELs, have been histopathologically validated as CALs, the lack of a correspondence/association between the two biomarkers suggests that SELs, as currently defined, may not reliably identify CALs *in vivo*.

### 4.4. Limitations

The principal limitation of this study lies in the relatively small sample size (56 subjects) and the specific characteristics of the cohort, a limitation driven by the conservative inclusion criteria for all participants to have a homogeneous advanced MRI protocol on the same 3T scanner for three consecutive timepoints. Notably, most PRLs in our dataset exhibited a reduction in volume over time, a pattern that diverges from trends commonly reported in the literature (Dal-Bianco et al., 2017, 2021; Weber et al., 2022), albeit in line with long-term observation (Sethi et al., 2017). Nonetheless, this does not affect the central conclusion: even if a biomarker demonstrates a consistent group-level trajectory, individual lesions can still display heterogeneous behaviors, with sub-groups of PRL in our cohort showing significantly different patterns of volumetric evolution. The limited cohort size also likely reduced statistical power, preventing the detection of a potential association between SEL volume with clinical disability score. Importantly, these limitations should not alter the broader conceptual issues of the SEL algorithm highlighted in the discussion.

## 5. Conclusion

In conclusion, our findings challenge the central assumptions underlying SELs: all CALs are not associated with volume expansion. Furthermore, the SEL detection algorithm lacks robustness against heterogeneous imaging acquisition and lesion segmentation, while the Jacobian-based methodology shows limited biological interpretability. The reliance on binary lesion masks instead of lesion-level instances, combined with population-dependent heuristic thresholds for SEL selection, further undermines the histopathological correspondence of this biomarker. Most critically, in validation experiments against a histopathologically-validated biomarker of CALs (i.e. PRLs), SELs demonstrated near complete independent correspondence, strongly suggesting that SELs, as currently defined, may not represent CALs.

Nevertheless, the longitudinal volumetric dynamics of MS lesions — whether expansion or shrinkage — remain of high interest, as they provide valuable insights into ongoing lesional activity *in vivo*. Future research should prioritize reproducible and open-source implementations that allow for independent verification and methodological transparency. Beyond expansion, lesion shrinkage should also be systematically evaluated, as both shrinking and expanding PRLs likely capture distinct aspects of chronic activity and tissue remodeling, with both showing evidence of longitudinal increased tissue damage. Furthermore, new methodological approaches should move away from heuristic population-based criteria and instead incorporate lesion-level features with stronger biological interpretability, possibly leveraging multi-contrast MRI. Finally, “MRI evolving lesions” could be conceptualized as a separate class of imaging biomarkers: informative about longitudinal dynamics and potentially relevant for disease monitoring, but distinct from histopathological lesion subtypes, whose longitudinal changes cannot be validated post-mortem.

## Supporting information

Automatic lesion splitting detailed in supplementary materials

## Data Availability

Data will be shared upon establishment of a data-sharing agreement with formal approval from the local ethics committee. All computational tools described in this study are openly released in the BMAT-Apps GitHub organization (https://github.com/orgs/BMAT-Apps) and can be freely downloaded and run using the BMAT Software (https://github.com/ColinVDB/BMAT).

## CRediT author statement

**CVB**: Data curation, Conceptualization, Methodology, Formal analysis, Investigation, Writing - Original Draft, Validation, Visualization

**AS**: Data Curation, Visualization, Formal analysis, Conceptualization, Writing - Review & Editing

**SB**: Data Curation, Visualization, Conceptualization, Writing - Review & Editing

**BM**: Writing - Review & Editing, Supervision, Resources, Funding acquisition

**MBC**: Writing – Review & Editing, Conceptualization, Investigation

**MA**: Writing – Review & Editing, Conceptualization, Investigation

**PM**: Writing - Original Draft, Conceptualization, Data Curation, Supervision, Investigation, Resources, Project administration

## Acknowledgements

The authors thank the study participants; the neuroimmunology clinics of cliniques universitaires Saint-Luc (CUSL, Brussels, Belgium) for recruiting and evaluating the patients and for coordinating the scans; Thierry Duprez, Sébastien de Laever (CUSL), Laurence Dricot (Université catholique de Louvain), Gaëtan Duchenes (CUSL), and Julie Poujol (GE Healthcare) for assistance with 3T MRI scan acquisition and analysis.

**CVB** is supported by the Fonds de Recherche Cliniques (FRC) from the Cliniques Universitaires Saint-Luc, UCLouvain, Brussels, Belgium, and by the MedReSyst-AI4Alzheimer project, which has been supported by the European Union and Wallonia as part of the “Wallonia 2021-2027” program.

**AS** has the financial support of the Fédération Wallonie Bruxelles – FRIA du Fonds de la Recherche Scientifique – FNRS.

**SB** is supported by the Funds Claire Fauconnier, Ginette Kryksztein & José and Marie Philippart-Hoffelt, managed by the King Baudouin Foundation.

**MBC** acknowledges the CIBM Center for Biomedical Imaging, a Swiss research center of excellence founded and supported by Lausanne University Hospital (CHUV), University of Lausanne (UNIL), École Polytechnique Fédérale de Lausanne (EPFL), University of Geneva (UNIGE) and Geneva University Hospitals (HUG).

**MA** received research support from the European Research Council (ERC), National MS Society, Conrad N Hilton Foundation, International Progressive MS Alliance, Cariplo Foundation, Fondazione Regionale Ricerca Biomedica (FRRB) Early Career Award, and FISM - Fondazione Italiana Sclerosi Multipla co-financed with the ‘5 per mille’ public funding.

**PM** research activity is supported by the Fondation Charcot Stichting Research Fund 2023, the Fund for Scientific Research (F.S.R, FNRS; grant 40008331), Cliniques universitaires Saint-Luc “Fonds de Recherche Clinique” and Biogen.

