## Supplementary material for "Do slowly expanding lesions correspond to chronic active multiple sclerosis lesions? An integrated imaging analysis study": Automatic lesion splitting detailed in supplementary materials

### Automatic lesion splitting

Lesion splitting is computed utilizing the fluid-attenuated inversion recovery (FLAIR) and magnetization prepared rapid gradient echo (MPRAGE) contrast to localize different clusters of tissue inside lesional area with different apparent T1w and T2w values, likely corresponding to different lesion instances with different ongoing pathological processes (representation of the lesion splitting in Figure S1).

First, preprocessing was applied on both FLAIR and MPRAGE images independently and included non-local means adaptive denoising filter (Manjón et al., 2010), skull-stripping using Synthstrip (Hoopes et al., 2022), rigid registration of the MPRAGE image onto the FLAIR space using the mutual information metric (ANTS) (Avants et al., 2008), and contrast enhancement using a voxel-wise squaring intensity transformation. Finally, we applied a frequency-domain sharpening step to both the MPRAGE and FLAIR images. The magnitude of their Fourier (F) transform was modulated via a power-law operator ($|F|^{0.75}$), and subsequently recombined with the unaltered phase for reconstruction. The inverse Fourier transform yielded images with enhanced high-frequency content, resulting in sharper lesion boundaries and facilitating subsequent separation of lesions.

Within the binary lesion mask obtained from FLAMeS on the FLAIR images (Dereskewicz et al., 2025), the sharpened FLAIR image was divided by the sharpened MPRAGE image to enhance contrast between regions with differing T2/T1 characteristics. The resulting map was further processed with a multi-scale Laplacian-of-Gaussian (LoG) filter to accentuate clusters with distinct T2/T1 ratios. Candidate lesion centers were identified as voxels where the gradient magnitude of the first derivative was near zero ($\varepsilon = 10^{-3}$) and where the Hessian's eigenvalues were all negative, indicating a local maximum. These centers were subsequently used as markers to initialize a watershed segmentation of the lesions, yielding the final lesion delineations.

To quantify the performance of the automated lesion-splitting approach, we carried out a correlation analysis comparing automated and manual lesion volumes. The evaluation was performed on a total of 1578 lesions manually delineated in the cross-sectional dataset. For each lesion, volumes derived from the automated algorithm were matched to their corresponding manual reference labels. Pearson's correlation was computed to quantify the performance of the automated lesion-splitting approach to truly represent the volume of individual lesions.

This evaluation revealed a robust correlation between automated and manual lesion volumes ($r = 0.8029$, 95% CI: [0.7845; 0.8197], $p < 0.00001$), indicating that the automated procedure reliably approximates expert-derived measurements (Figure S2).

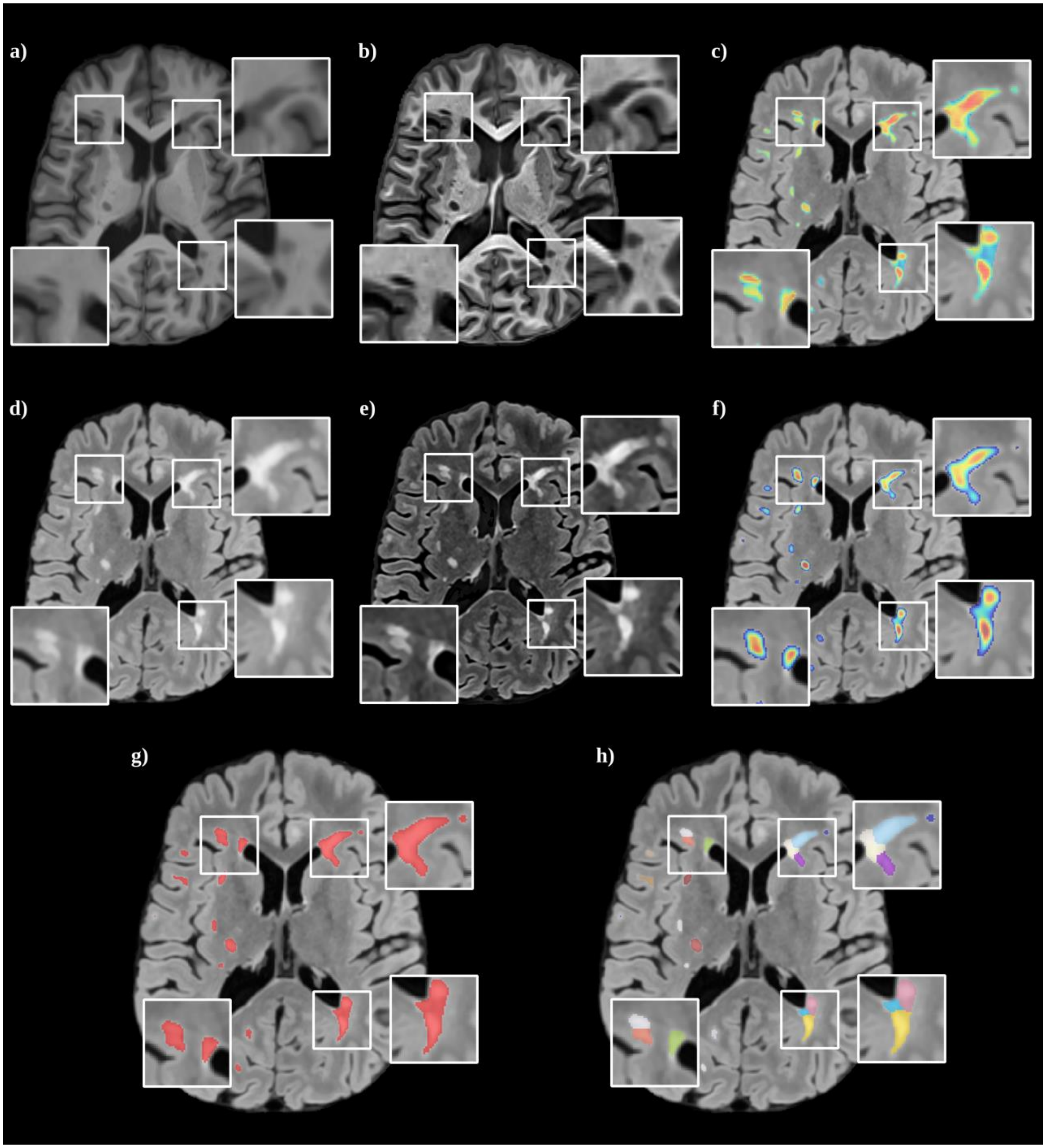


**Figure S1.** Representative figure of the lesion-splitting algorithm in axial images of an individual in their 20’s with relapsing-remitting multiple sclerosis (RRMS). (a) magnetization prepared rapid gradient echo (MPRAGE) image; (b) preprocessed and sharpened MPRAGE image; (c) fluid-attenuated inversion recovery (FLAIR) image with the FLAIR/MPRAGE ratio overlay; (d) FLAIR image; (e) preprocessed and sharpened FLAIR image; (f) FLAIR image with the multi-scales Laplacian of gaussian (LoG) filter applied on the FLAIR/MPRAGE ratio overlay; (g) FLAIR image with automatic binary lesion segmentation; (h) FLAIR image with the resulting lesion-splitted segmentation.

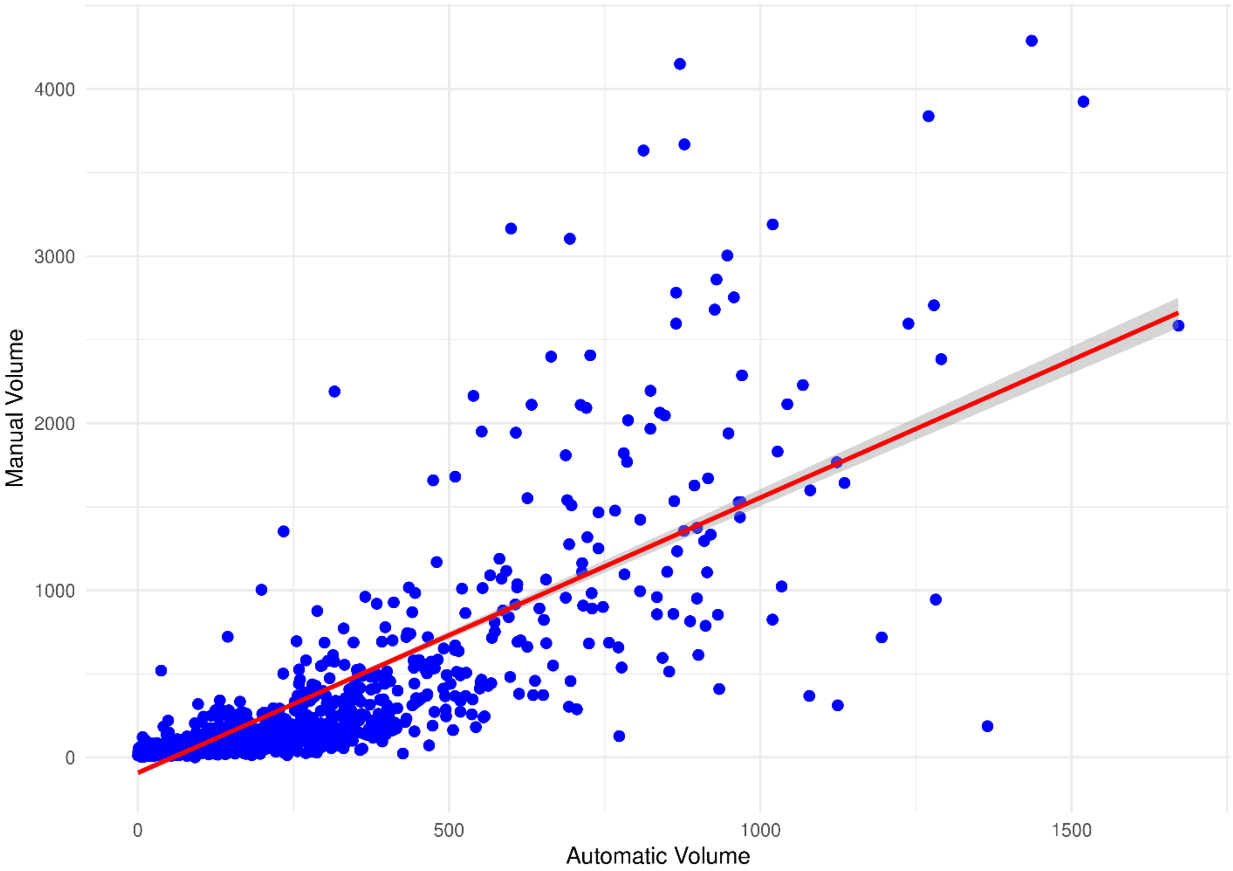


**Figure S2.** Scatterplot illustrating the relationship between automated and manual lesion volumes. The line of best fit and corresponding correlation coefficient ($r = 0.8029$, 95% CI: [0.7845; 0.8197], $p < 0.00001$) indicates strong agreement between the two methods.
